## Supplementary material for "Distinguishing non severe cases of dengue from COVID-19 in the context of co-epidemics: a cohort study in a SARS-CoV-2 testing center on Reunion island": Methodological appendix

**Methods**

**Study design, setting and population**

We conducted a retrospective cohort study using prospectively collected data between March 23 and May 10, 2020, on all subjects screened for the COVID-19 within the UDACS (*Unité de Dépistage Ambulatoire du COVID-19 Sud*) of Saint-Pierre, one of the two SARS-CoV-2 testing centers of the *Centre Hospitalier Universitaire* (CHU) *Réunion*. When SARS-CoV-2 emerged on the island, the dengue epidemic was already burgeoning, the UDACS was placed in the second line of the reception system for COVID-19 patients, the frontline being the emergency units and the dedicated hospital for COVID-19 patients, the CHU Félix Guyon, located in Saint-Denis, whereby are the prefecture and the international airport. People without symptoms were excluded from the study.

**Ethics statement**

Outpatients presenting consecutively at the SARS-CoV-2 testing center were informed of the study orally and by means of an information sheet. Adult people, like the children under 18 years (with the additional verbal consent of their parent or legal guardian) who expressed no opposition, were asked to answer a questionnaire and surveyed in face-to-face by a nurse, in accordance to the French legislation on bioethics for retrospective researches. Patient’s medical records were retrospectively reviewed, and de-identified data were collected in standardized forms according to the MR-004 procedure of the *Commission Nationale de l’Informatique et des Libertés* (the French information protection commission). The ethical character of this study on previously collected data was approved by the Scientific Committee for COVID-19 research of the CHU Réunion and de-identified data were registered on the Health Data Hub.

**Data collection**

The items of the questionnaire included information on demographics (gender, age), occupation, risk factors (smoking, obesity, recent travel abroad < 15 days), comorbidities (diabetes, hypertension, cardiovascular disease, chronic obstructive pulmonary disease, cancer, previous episode of dengue, other), intra-household and individual exposure to SARS-CoV-2, individual symptoms (fever, cough, dyspnea/shortness of breath, body ache, diarrhea, nausea, vomiting, dyspepsia, eructation, abdominal pain, ageusia, dysgeusia/metallic taste, anosmia, fatigue, headache, retro-orbital pain, sore throat, runny nose, nasal congestion, sneezing) and treatment (antihypertensive drugs, hydroxychloroquine). Temperature, pulse rate, respiratory rate and oxygen saturation (SpO_2_) were measured upon the consultation, as well as the presence of cough and anxiety. People reporting symptoms were physically examined by a resident and further by a senior infectious disease doctor, according to routine care procedures.

**Diagnostic procedures**

All the attendees were screened by a skilled nurse for SARS-CoV-2 using a nasopharyngeal swab inserted and held in one nostril until reaching the posterior wall of the nasopharynx for about twenty seconds [1]. The sample was processed for a SARS-CoV-2 reverse transcription-polymerase chain reaction (RT-PCR) using a Microlab NIMBUS/STARlet IVD (Seegene, Seoul, Republic of Korea) or a MagNa Pure Compact® (Roche Life Science, Penzberg, Germany) automaton for nucleic acid extraction. SARS-CoV-2 ribonucleic acid sequences were identified using the Allplex 2019-nCov^TM^ assay (Seegene, Seoul, Republic of Korea) or an in-house kit (CNR Pasteur), targeting N, RdRP and E genes, or N and IP2/IP4 targets of RdRP, respectively. SARS-CoV-2 genome sequences were amplified using a Bio-Rad CFX96® (BioRad, Hercules, CA, USA) or a LightCycler 480®(Roche Diagnostics, Basel, Switzerland) system. In addition, each patient suspected of dengue was tested for NS1 antigen using an OnSite^TM^ Duo dengue Ag-IgG-IgM rapid diagnostic test (CTK Biotech, San Diego, CA, USA) and if negative further explored with a DENV RT-PCR (NucliSens easyMAG, Biomerieux®, France; LightCycler 480®, Roche Dagnostics, Basel, Switzerland) or a Panbio® dengue Duo (IgM/IgG) capture ELISA (Abbott, Chicago, IL, USA) depending on the date of symptom onset. Patients requiring hospitalization were transferred promptly from the UDACS to the COVID-19 units. Patients not requiring hospitalization were discharged home and benefited an ambulatory follow-up ensured by general practitioners.

**Statistical analysis**

Given the research purpose, COVID-19-dengue co-infections at clinical presentation were excluded from the analysis. Other febrile illnesses (OFIs) were defined as patients tested negative for SARS-CoV-2 and further considered as unrelated to dengue, either clinically, virologically, or serologically. Proportions between non COVID-19 and non-dengue other febrile illnesses (OFIs), COVID-19 and dengue subjects were compared using Chi square or Fisher exact tests, as appropriate.

Univariable and multivariable multinomial logistic regression models were fitted within Stata14® (StataCorp, College Station, Texas, USA) to identify both the independent predictors of COVID-19 and dengue, taking OFIs as controls. At the first step, we fitted a full multinomial logistic regression model with all significant variables identified by univariable analysis. From these covariates, we used a backward stepwise selection procedure to drop out the non- significant variables (output if *P* value >0.05). At third step, we built a minimal multinomial logistic regression model with all significant covariates from the precedent model.

We next performed weighted analyses on the overall inverse probability of hospitalization to assess the potential for selection bias. Briefly, we applied in logistic regression models the hospitalization rates observed for confirmed COVID-19 and dengue cases over the study period, the actual rate (1.5%) or a probability 16% for OFIs using the *svy* function in Stata.

To limit the potential for misclassification bias in the differential diagnosis between COVID-19 and dengue due to the overlapping symptoms between COVID-19 and OFIs, we also performed a sensitivity analysis restricted to confirmed COVID-19 and dengue cases *(i.e.*, excluding OFIs).

Crude and adjusted odds ratios (OR) and 95% confidence intervals (95%CI) were assessed using the binomial and Cornfield methods, respectively. All interaction terms between the variables were tested. The log-likelihood of the models was tested together with the maximum likelihood test, the Mac-Fadden pseudo R^2^ and the Bayesian Information criterion (BIC) measures within the *fitstat* module [2]. The calibration of the models (*i.e.*, the adequacy between predicted cases and observed cases) was evaluated using the modified Hosmer and Lemeshow goodness-of-fit test for multinomial logistic regression with the *mlogitgof* command [3]. The discriminative power of the models for the diagnosis of COVID-19 and dengue was tested using the C-statistic which displays areas under the receiver operating characteristics curves (AuROC) with their 95%CIs.

For all these analyses, observations with missing data were ruled out and a *P*-value less than 0.05 was considered as statistically significant.
