## Supplementary tables for "Distinguishing non severe cases of dengue from COVID-19 in the context of co-epidemics: a cohort study in a SARS-CoV-2 testing center on Reunion island"

**Table S1. Crude predictors in bivariate analysis distinguishing COVID-19 and dengue from other febrile illnesses among 1,013 subjects consulting a COVID-19 screening center during the COVID-19 dengue co-epidemics, Reunion island, Saint-Pierre, March 23-May 10, 2020**

| Outcomes (vs other febrile illnesses as controls*) | COVID-19 (n = 80) | | | | | Dengue (n = 61) | | | | |
| --- | --- | --- | --- | --- | --- | --- | --- | --- | --- | --- |
| Predictors | **n** | **CIR, %** | **cOR** | **95% CI** | ***P* value** | **n** | **CIR, %** | **cOR** | **95% CI** | ***P* value** |
| Gender |  |  |  |  |  |  |  |  |  |  |
| Male | 33 | 8.11 | 1 |  |  | 31 | 7.62 | 1 |  |  |
| Female | 47 | 7.76 | 0.92 | 0.57 - 1.47 | 0.738 | 30 | 4.95 | 0.63 | 0.37 - 1.06 | 0.079 |
| Age, years |  |  |  |  |  |  |  |  |  |  |
| 0-30 (Q1) | 28 | 9.69 | 1.11 | 0.57 - 2.15 | 0.748 | 8 | 2.77 | 0.34 | 0.13 - 0.83 | 0.018 |
| 31-41 (Q2) | 10 | 3.53 | 0.41 | 0.17 - 0.93 | 0.033 | 26 | 9.19 | 1.13 | 0.57 - 2.24 | 0.715 |
| 42-54 (Q3) | 27 | 10.34 | 1.23 | 0.63 - 2.39 | 0.542 | 13 | 4.98 | 0.63 | 0.29 - 1.39 | 0.255 |
| 55-94 (Q4) | 15 | 8.33 | 1 |  |  | 14 | 7.78 | 1 |  |  |
| Contact with a COVID-19 positive case |  |  |  |  |  |  |  |  |  |  |
| No | 38 | 5.18 | 1 |  |  | 55 | 7.49 | 1 |  |  |
| Yes | 42 | 15.05 | 3.07 | 1.22 - 4.88 | < 0.001 | 6 | 2.15 | 0.30 | 0.12 - 0.71 | 0.006 |
| Return from travel abroad < 15 days |  |  |  |  |  |  |  |  |  |  |
| No | 37 | 4.87 | 1 |  |  | 55 | 7.24 | 1 |  |  |
| Yes | 42 | 16.80 | 3.75 | 2.34 - 6.00 | < 0.001 | 6 | 2.40 | 0.36 | 0.15 - 0.85 | 0.020 |
| Previous dengue episode |  |  |  |  |  |  |  |  |  |  |
| No | 73 | 7.56 | 1 |  |  | 52 | 5.39 | 1 |  |  |
| Yes | 6 | 12.77 | 2.16 | 0.87 - 5.33 | 0.096 | 9 | 19.15 | 4.54 | 2.05 - 10.02 | < 0.001 |
| Active smoking ^†^ |  |  |  |  |  |  |  |  |  |  |
| No | 73 | 8.63 | 1 |  |  | 49 | 5.79 | 1 |  |  |
| Yes | 4 | 2.47 | 0.27 | 0.09 - 0.76 | 0.015 | 12 | 7.41 | 1.21 | 0.63 - 2.34 | 0.562 |
| Fever |  |  |  |  |  |  |  |  |  |  |
| No | 3 | 6.55 | 1 |  |  | 2 | 0.37 | 1 |  |  |
| Yes | 45 | 9.41 | 1.71 | 1.07 - 2.71 | 0.023 | 59 | 12.34 | 39.20 | 9.51 - 161.57 | < 0.001 |
| Cough |  |  |  |  |  |  |  |  |  |  |
| No | 44 | 8.38 | 1 |  |  | 44 | 8.38 | 1 |  |  |
| Yes | 36 | 7.38 | 1 | 0.51 - 1.30 | 0.404 | 17 | 3.48 | 0.39 | 0.21 - 0.69 | 0.001 |
| Dyspnea/Shortness of breath |  |  |  |  |  |  |  |  |  |  |
| No | 67 | 8.56 | 1 |  |  | 48 | 6.13 | 1 |  |  |
| Yes | 13 | 5.65 | 0.64 | 0.34 - 1.17 | 0.148 | 13 | 5.65 | 0.89 | 0.47 - 1.67 | 0.710 |
| Body ache ^‡^ |  |  |  |  |  |  |  |  |  |  |
| No | 48 | 8.14 | 1 |  |  | 9 | 1.53 | 1 |  |  |
| Yes | 32 | 7.57 | 1.05 | 0.65 - 1.67 | 0.844 | 52 | 12.29 | 9.08 | 4.41 - 18.68 | < 0.001 |
| Diarrhea |  |  |  |  |  |  |  |  |  |  |
| No | 61 | 7.59 | 1 |  |  | 48 | 5.97 | 1 |  |  |
| Yes | 19 | 9.13 | 1.23 | 0.71 - 2.11 | 0.453 | 13 | 6.25 | 1.07 | 0.56 - 2.02 | 0.836 |
| To be continued… |  |  |  |  |  |  |  |  |  |  |
| Gut symptoms ^¶^ |  |  |  |  |  |  |  |  |  |  |
| No | 76 | 7.98 | 1 |  |  | 48 | 5.04 | 1 |  |  |
| Yes | 4 | 6.56 | 0.99 | 0.34 - 2.83 | 0.986 | 13 | 21.31 | 5.10 | 2.57 - 10.10 | < 0.001 |
| Ageusia |  |  |  |  |  |  |  |  |  |  |
| No | 55 | 6.16 | 1 |  |  | 50 | 5.60 | 1 |  |  |
| Yes | 25 | 20.83 | 4.26 | 2.52 - 7.19 | < 0.001 | 11 | 9.17 | 2.06 | 1.03 - 4.12 | 0.040 |
| Metallic taste (dysgeusia) |  |  |  |  |  |  |  |  |  |  |
| No | 80 | 7.94 | 1 |  |  | 59 | 5.86 | 1 |  |  |
| Yes | 0 | 0.00 | N.A |  | 0.544 | 2 | 33.3 | 7.36 | 1.32 - 40.99 | 0.008 |
| Anosmia |  |  |  |  |  |  |  |  |  |  |
| No | 52 | 5.68 | 1 |  |  | 58 | 6.34 | 1 |  |  |
| Yes | 28 | 28.57 | 6.47 | 3.83 - 10.91 | < 0.001 | 3 | 3.06 | 0.62 | 0.18 - 2.04 | 0.432 |
| Fatigue |  |  |  |  |  |  |  |  |  |  |
| No | 42 | 7.55 | 1 |  |  | 12 | 2.16 | 1 |  |  |
| Yes | 38 | 8.32 | 1.23 | 0.77 - 1.94 | 0.381 | 49 | 10.72 | 5.54 | 2.90 - 10.57 | < 0.001 |
| Headache |  |  |  |  |  |  |  |  |  |  |
| No | 49 | 9.51 | 1 |  |  | 5 | 0.97 | 1 |  |  |
| Yes | 31 | 6.24 | 0.71 | 0.44 - 1.14 | 0.155 | 56 | 11.27 | 12.59 | 4.99 - 31.75 | < 0.001 |
| Retro-orbital pain |  |  |  |  |  |  |  |  |  |  |
| No | 79 | 8.15 | 1 |  |  | 44 | 4.54 | 1 |  |  |
| Yes | 1 | 2.27 | 0.41 | 0.05 - 3.08 | 0.401 | 17 | 38.64 | 12.57 | 6.35 - 24.88 | < 0.001 |
| URTI symptoms ^#^ |  |  |  |  |  |  |  |  |  |  |
| No | 49 | 9.74 | 1 |  |  | 41 | 8.15 | 1 |  |  |
| Yes | 31 | 6.08 | 0.57 | 0.35 - 0.91 | 0.019 | 20 | 3.92 | 0.44 | 0.25 - 0.76 | 0.003 |
| Presentation > 3 days after symptom onset |  |  |  |  |  |  |  |  |  |  |
| No | 23 | 5.49 | 1 |  |  | 36 | 8.59 | 1 |  |  |
| Yes | 54 | 9.66 | 1.76 | 1.05 - 2.92 | 0.029 | 24 | 4.29 | 0.50 | 0.29 - 0.85 | 0.011 |
| * Other non COVID-19 non dengue febrile illnesses. Data are numbers, cumulative incidence rates (CIR) expressed as percentages, crude odd ratios (cOR), 95% confidence intervals (95% CI) and *P* values for Wald tests. † Current smokers, as compared to never smokers and past smokers ^‡^ muscle pain or backache with tightness and/or stiffness; ^¶^ nausea, vomiting, dyspepsia, eructation or abdominal pain ^#^ sore throat, runny nose, nasal congestion, or sneezing. N.A: not assessed (incalculable). | | | | | | | | | | |

**Table S2. Sensitivity analysis. Crude predictors in bivariate analysis distinguishing COVID-19 from dengue from after exclusion of other febrile illnesses among 141 subjects consulting a COVID-19 screening center during the COVID-19 dengue co-epidemics, Reunion island, Saint-Pierre, March 23-May 10, 2020**

| Outcomes | COVID-19  (n = 80) | | Dengue  (n = 61) | |  |
| --- | --- | --- | --- | --- | --- |
| Predictors | **n** | **(%)** | **n** | **(%)** | ***P* value** |
| Gender |  |  |  |  | 0.258 |
| Male | 33 | 51.6 | 31 | 48.4 |  |
| Female | 47 | 61.0 | 30 | 39.0 |  |
| Age, years |  |  |  |  | < 0.001 |
| 0-30 (Q1) | 28 | 77.8 | 8 | 22.2 |  |
| 31-41 (Q2) | 10 | 27.8 | 26 | 72.2 |  |
| 42-54 (Q3) | 27 | 67.5 | 13 | 32.5 |  |
| 55-94 (Q4) | 15 | 51.7 | 14 | 48.3 |  |
| Contact with a COVID-19 positive case |  |  |  |  | < 0.001 |
| No | 38 | 40.9 | 55 | 59.1 |  |
| Yes | 42 | 87.5 | 6 | 12.5 |  |
| Return from travel abroad < 15 days |  |  |  |  | < 0.001 |
| No | 37 | 40.2 | 55 | 59.8 |  |
| Yes | 42 | 87.5 | 6 | 12.5 |  |
| Previous dengue episode |  |  |  |  | 0.174 |
| No | 73 | 58.4 | 52 | 41.6 |  |
| Yes | 6 | 40.0 | 9 | 60.0 |  |
| Active smoking ^†^ |  |  |  |  | 0.008 |
| No | 73 | 59.8 | 49 | 40.2 |  |
| Yes | 4 | 25.0 | 12 | 75.0 |  |
| Fever |  |  |  |  | < 0.001 |
| No | 35 | 94.6 | 2 | 5.4 |  |
| Yes | 45 | 43.3 | 59 | 56.7 |  |
| Duration of fever (days), µ ± sd | 3.43 | 3.35 | 3.03 | 2.88 | 0.892 |
| Cough |  |  |  |  | 0.037 |
| No | 44 | 50.0 | 44 | 50.0 |  |
| Yes | 36 | 67.9 | 17 | 32.1 |  |
| Duration of cough (days), µ ± sd | 2.14 | 12.84 | 5.79 | 7.98 | 0.958 |
| Dyspnea/Shortness of breath |  |  |  |  | 0.443 |
| No | 67 | 58.3 | 48 | 41.7 |  |
| Yes | 13 | 50.0 | 13 | 50.0 |  |
| Duration of dyspnea (days), µ ± sd | 5.44 | 8.43 | 7.75 | 5.25 | 0.771 |
| Body ache ^‡^ |  |  |  |  | < 0.001 |
| No | 48 | 84.2 | 9 | 15.8 |  |
| Yes | 32 | 38.1 | 52 | 61.9 |  |
| Duration of pain (days), µ ± sd | 4.34 | 5.49 | 2.90 | 2.72 | 0.399 |
| Diarrhea |  |  |  |  | 0.732 |
| No | 61 | 56.0 | 48 | 44.0 |  |
| Yes | 19 | 59.4 | 13 | 40.6 |  |
| Duration of liquid stools (days), µ ± sd | 4.50 | 3.79 | 2.25 | 3.14 | 0.087 |
| Gut symptoms ^¶^ |  |  |  |  | 0.003 |
| No | 76 | 61.3 | 48 | 38.7 |  |
| Yes | 4 | 23.5 | 13 | 76.5 |  |
| Ageusia |  |  |  |  | 0.075 |
| No | 55 | 52.4 | 50 | 47.6 |  |
| Yes | 25 | 69.4 | 11 | 30.6 |  |
| Duration of ageusia (days), µ ± sd | 4.73 | 3.32 | 3.25 | 2.01 | 0.271 |
| To be continued… |  |  |  |  |  |
| Anosmia |  |  |  |  | < 0.001 |
| No | 52 | 47.3 | 58 | 52.7 |  |
| Yes | 28 | 90.3 | 3 | 9.7 |  |
| Duration of anosmia (days), µ ± sd | 4.22 | 3.59 | 1.00 | 1.00 | 0.093 |
| Fatigue |  |  |  |  | < 0.001 |
| No | 42 | 77.8 | 12 | 22.2 |  |
| Yes | 38 | 43.7 | 49 | 56.3 |  |
| Duration of fatigue (days), µ ± sd | 6.48 | 5.75 | 3.44 | 3.00 | 0.009 |
| Headache |  |  |  |  | < 0.001 |
| No | 49 | 90.7 | 5 | 9.3 |  |
| Yes | 31 | 35.6 | 56 | 64.4 |  |
| Duration of headache (days), µ ± sd | 4.69 | 5.61 | 3.02 | 2.74 | 0.216 |
| Retro-orbital pain |  |  |  |  | < 0.001 |
| No | 79 | 64.2 | 44 | 35.8 |  |
| Yes | 1 | 5.6 | 17 | 94.4 |  |
| URTI symptoms ^#^ |  |  |  |  | 0.465 |
| No | 49 | 54.4 | 41 | 45.6 |  |
| Yes | 31 | 60.8 | 20 | 39.2 |  |
| Duration of rhinorrhea (days), µ ± sd | 5.33 | 3.69 | 2.10 | 0.91 | 0.008 |
| Presentation > 3 days after symptom onset |  |  |  |  | < 0.001 |
| No | 23 | 39.0 | 36 | 61.0 |  |
| Yes | 54 | 69.2 | 24 | 30.8 |  |
| Time elapsed since symptom onset (days), µ ± sd | 7.54 | 6.50 | 4.18 | 4.57 | < 0.001 |
| * Other non COVID-19 non dengue febrile illnesses. Data are numbers, row percentages, and *P* values for Chi2 or Fisher’s exact tests, unless specified as means, standard deviations, and *P* values for Mann-Whitney tests. † Current smokers, as compared to never smokers and past smokers ^‡^ muscle pain or backache with tightness and/or stiffness; ^¶^ nausea, vomiting, dyspepsia, eructation or abdominal pain ^#^ sore throat, runny nose, nasal congestion, or sneezing. | | | | | |
