## Supplementary figures and images for "Distinguishing non severe cases of dengue from COVID-19 in the context of co-epidemics: a cohort study in a SARS-CoV-2 testing center on Reunion island"

### Supplementary figure

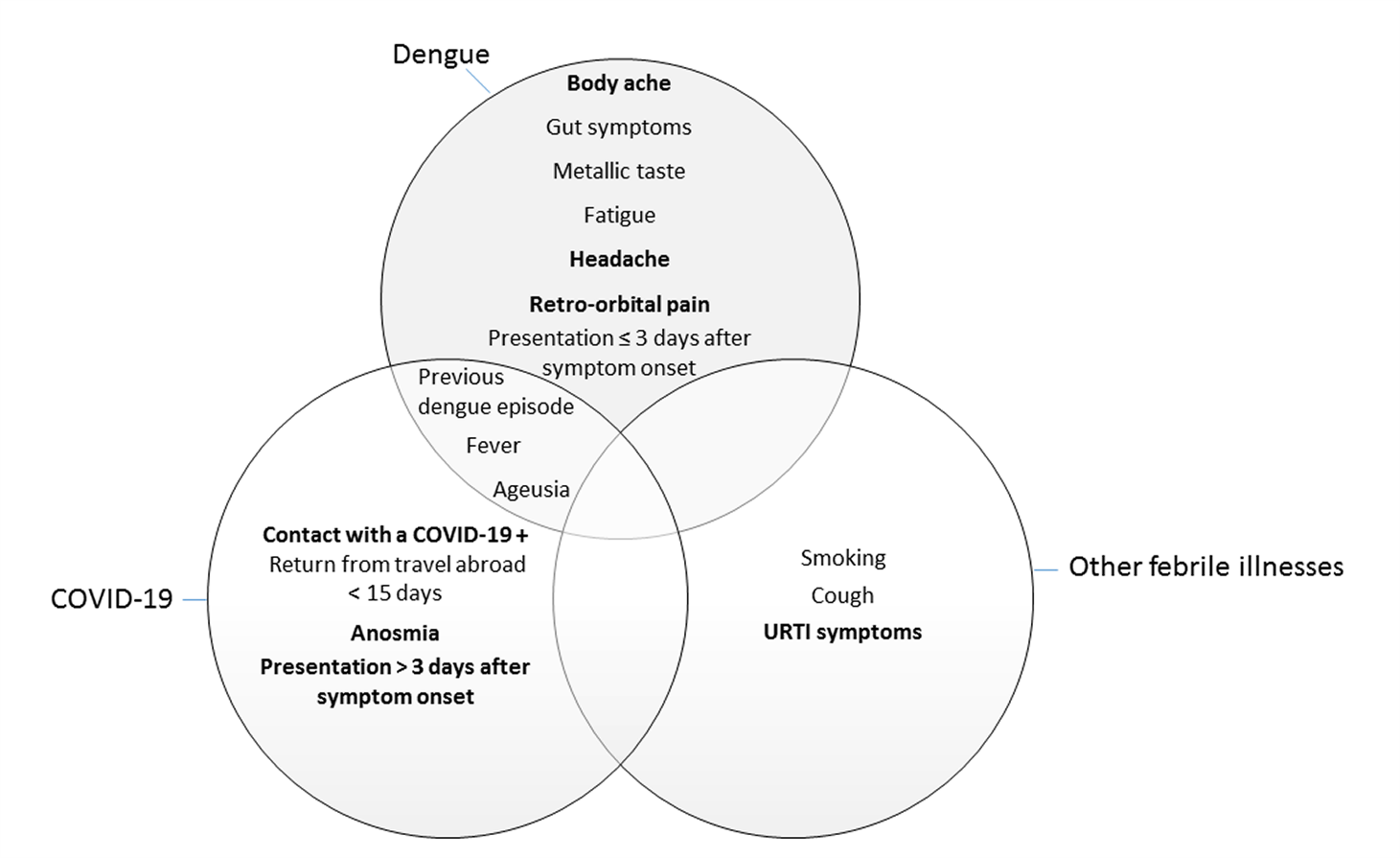
